# Impact and cost-effectiveness of measles vaccination through microarray patches in 70 low- and middle-income countries: a modelling study

**DOI:** 10.1101/2023.03.10.23287067

**Authors:** Han Fu, Kaja Abbas, Stefano Malvolti, Christopher Gregory, Melissa Ko, Jean-Pierre Amorij, Mark Jit

## Abstract

**Background:** Microarray patches are a promising technology being developed to reduce barriers to vaccine delivery based on needles and syringes. To address the evidence gap on the public health value of applying this potential technology to immunisation programmes, we evaluated the health impact on measles burden and resulting cost-effectiveness of introducing measles-rubella microarray patches (MR-MAPs) in 70 low- and middle-income countries (LMICs).

**Methods:** We used an age-structured dynamic model of measles transmission and vaccination to project measles cases, deaths, and disability-adjusted life years during 2030⍰2040. Compared to the baseline scenarios with continuing current needle-based immunisation practice, we evaluated the introduction of MR-MAPs under different assumptions on measles vaccine coverage projections and MR-MAP introduction strategies. Costs were calculated based on the ingredients approach, including direct cost of measles treatment, vaccine procurement, and vaccine delivery. Model-based burden and cost estimates were derived for individual countries and country income groups. We compared the incremental cost-effectiveness ratios of introducing MR-MAPs to health opportunity costs.

**Results:** MR-MAPs introduction could prevent 27%⍰37% of measles burden between 2030⍰2040 in 70 LMICs. The largest health impact could be achieved under lower coverage projection and accelerated introduction strategy, with 39 million measles cases averted. Cost of measles treatment is a key driver of the net cost of introduction. In LMICs with a relatively higher income, introducing MR-MAPs could be a cost-saving intervention due to reduction in measles treatment costs. Compared to country-specific health opportunity costs, introducing MR-MAPs would be cost-effective in 16⍰81% of LMICs, depending on the MR-MAPs procurement price and vaccine coverage projections.

**Conclusions:** Introducing MR-MAPs in LMICs can be a cost-effective strategy to revitalise measles immunisation programmes with stagnant uptake and reach under-vaccinated children. Sustainable introduction and uptake of MR-MAPs has the potential to improve vaccine equity within and between countries and accelerate progress towards measles elimination.

## Introduction

As one of the essential childhood immunisations, measles-containing vaccines (MCV) has been recommended by the World Health Organization (WHO) Expanded Programme on Immunisation since 1974.^1^ The implementation of measles vaccination has brought substantial health benefits, with an estimated 33 million deaths averted between 2000⍰2019.^2^ Maintaining high coverage (>95%) with two doses of MCV is also recognised as a core strategy for measles elimination.^3^ However, the uptake of the first dose of MCV was stagnant at 85% globally over 2011⍰2019, while measles outbreaks continued to occur, and measles remained a major public health burden in settings with low MCV coverage. The COVID-19 pandemic posed further challenges to national immunisation programmes, as seen in a 5% drop in the global coverage of first-dose MCV from 2019 to 2021.^4 5^ The coverage of the second MCV dose has steadily increased and reached 72% in 2020, but its progress varies across countries and has stagnated in the last few years.^5^

Currently, MCVs are delivered via needles and syringes (N&Ss). The traditional N&S presentation requires reconstitution before administration, and reconstituted doses that are not used within six hours are discarded. The delivery of N&S vaccines relies on trained healthcare workers for administration and demands a comprehensive and well-functioning cold chain system for storage and transportation. Addressing barriers to effective vaccine delivery associated with N&S vaccines could reduce global measles burden and accelerate progress in measles elimination.

Microarray patches (MAPs), a device containing hundreds to thousands of micro-projections that deliver a vaccine dose into the dermis, has product characteristics that could address the barriers to vaccination presented by N&Ss. MAPs have demonstrated stability under higher temperatures for several vaccines, which reduces cold chain demand and potentially makes it easier to deliver vaccines to hard-to-reach areas.^6 7^ In the absence of needles, injection applicators, and reconstitution devices, MAPs could be operated by minimally trained staff or self-administrated, which may expand the workforce to reach zero-dose children and under-immunised population.^8^ MAPs are also broadly applicable in routine immunisation (RI) programmes and supplementary immunisation activities (SIAs). In low- and middle-income countries (LMICs), implementing MAP technology may transform the current delivery of measles and rubella vaccines in immunisation programmes and better reach under-served populations in remote rural or conflict-affected areas.^9-11^ In May 2020, the Vaccine Innovation Prioritisation Strategy, a three-year collaboration between Gavi, the Vaccine Alliance, WHO, Bill & Melinda Gates Foundation, United Nations Children’s Fund (UNICEF) and PATH to develop a single integrated framework to evaluate, prioritise, and drive forwards vaccine product innovations, selected MAPs as one of three technologies for prioritisation for development and implementation.^8^

As measles and rubella vaccines are jointly administered in most settings, a bivalent measles-rubella MAP (MR-MAP) is considered to have significantly broader use than a monovalent measles MAP.^9^ MR-MAPs demonstrated both immunogenicity and safety in preclinical studies in infant rhesus macaques and provided effective protection against wild-type measles challenge.^12^ Ongoing Phase I/II trials of MR-MAPs are conducted in The Gambia^13^ and Australia.^14^ Despite the early stage of clinical development, economic evaluation for MR-MAPs prior to phase III trials helps to determine the public health impact and economic case for further investment in development, as well as identify sources of uncertainties around potential impact and cost-effectiveness to inform directions of data collection in the future.^15^ Furthermore, early-stage economic evaluation can assess key determinants of cost-effectiveness and provide feedback on the MR-MAP product profile.^16^

In 2021, UNICEF commissioned MMGH Consulting GmbH in partnership with London School of Hygiene and Tropical Medicine and Global Health Visions to conduct an initial Full Value of Vaccine Assessment for MR-MAPs, to improve the assessment, decision-making, and communication concerning MR-MAP development, procurement, and implementation particularly for the use in LMICs. Placing end-users and stakeholders at the centre, the Assessment aims to facilitate discussion and coordination between different perspectives and address various impacts on health, economics, and society.^17^ Analyses to understand the comprehensive value of MR-MAPs, including the total system costs, commercial business case, and needs for market incentives, are available in the report for the (initial) Full Value of Vaccines Assessment of MR-MAPs (*DOI will be added once officially released*.). As part of the value assessment framework for MR-MAPs, this analysis focused on the health impact and cost-effectiveness of introducing MR-MAPs in LMICs, where more than 90% of global measles cases occur.^5^

## Methods

### Study setting

We included 70 LMICs in this global analysis of measles burden and vaccination, including 20 low-income countries, 35 lower-middle-income countries, and 15 upper-middle-income countries based on the World Bank income classification for the 2022 fiscal year.^18^ We included all LMICs apart from those having been verified for measles elimination and then kept their status until at least 2019 and those LMICs having ≥ 95% coverage with two doses of MCV and reporting ≤ 5 annual measles cases during 2017–2019. Additionally, we excluded countries that were projected to achieve a low level of measles burden in our analysis (see ‘Coverage Forecasts’ section), defined as ≤ 5 annual cases over 2027–2029 and thus for which introducing MR-MAPs after 2030 would be expected to have only marginal benefit in burden reduction (Albania, Botswana, Cape Verde, Djibouti, Eritrea, Fiji, Georgia, Guinea, Guinea-Bissau, Jordan, Kazakhstan, Kuwait, Kyrgyzstan, Malaysia, Morocco, Mauritius, Rwanda, South Sudan, Thailand, Tonga, and Vanuatu). The full list of countries included in the analysis can be found in table A in online supplemental file S1.

### Epidemiological model

We assessed the epidemiological impact of MR-MAPs introduction on measles burden in each country using Dynamic Measles Immunisation Calculation Engine (DynaMICE), for which a list of key parameters are included in table B in online supplemental file S1 and a detailed description and application has been published.^19^ DynaMICE is an age-structured compartmental model of measles transmission with states for people who are susceptible, infectious, recovered, protected by maternal antibodies, and protected by immunisation. Measles transmission between age groups is simulated using country-specific social contact matrices^20^ with a basic reproduction number of 15.9 based on a systematic review.^21^ Case-fatality risks of measles are varied across countries and are relatively higher for children under 5 years old based on a recent review of available data.^22^

DynaMICE takes into account MCV coverage, efficacy, age at vaccination, and vaccination history of targeted populations. In the model, RI programmes provide the first (MCV1) and second (MCV2) doses of measles vaccines to children aged 9 months and 16.5 months, respectively. SIAs are regularly scheduled for a population of a selected age range to enhance MCV coverage. We assumed SIA doses are delivered to children regardless of their vaccination history, except for up to 7.7% of children who are unreachable under current vaccination activities.^23^ In the model, the efficacy of the first MCV dose increases linearly with the age of vaccine administration^24^ and receiving two MCV doses provides 98% efficacy.^25^ Vaccine protection was assumed to be all-or-nothing, i.e., to offer complete protection to a proportion of the vaccinated individuals and no protection to the rest.

### Coverage projection

Aligning with the approach used in Global Market Study for MCVs,^26^ Ko et al. projected global demand for MCV doses over 2030⍰2040 and developed use cases of MR-MAPs in needle-based immunisation programmes at the country level.^27^ The forecasts were adapted to coverage inputs and delivery components for the scenarios we evaluated in this cost-effectiveness analysis of introducing MR-MAPs.

To capture uncertainty in the future uptake of measles vaccination, especially given that countries are rebuilding health services disrupted by the COVID-19 pandemic, ‘higher’ and ‘lower’ coverage projections for MCV coverage were considered (table 1). Under the ‘higher’ coverage projection, current RI coverage for MCV1 and MCV2 is projected to increase by 0.5⍰3% per year and capped at 95% or a higher level seen in the country-specific historical coverage; SIAs will take place every 2⍰5 years with a 95% coverage of children aged 9⍰59 months and will discontinue when MCV2 coverage exceeds 90% over 3 consecutive years. The growth rate of RI coverage and frequency of SIAs depend on country immunisation programmes. Under the ‘lower’ coverage projection, where a less optimistic perspective is applied to the future programme expansion, projected RI coverage will remain constant at the 2019 level, and SIAs will reach only 85% of children.

**Table 1.**
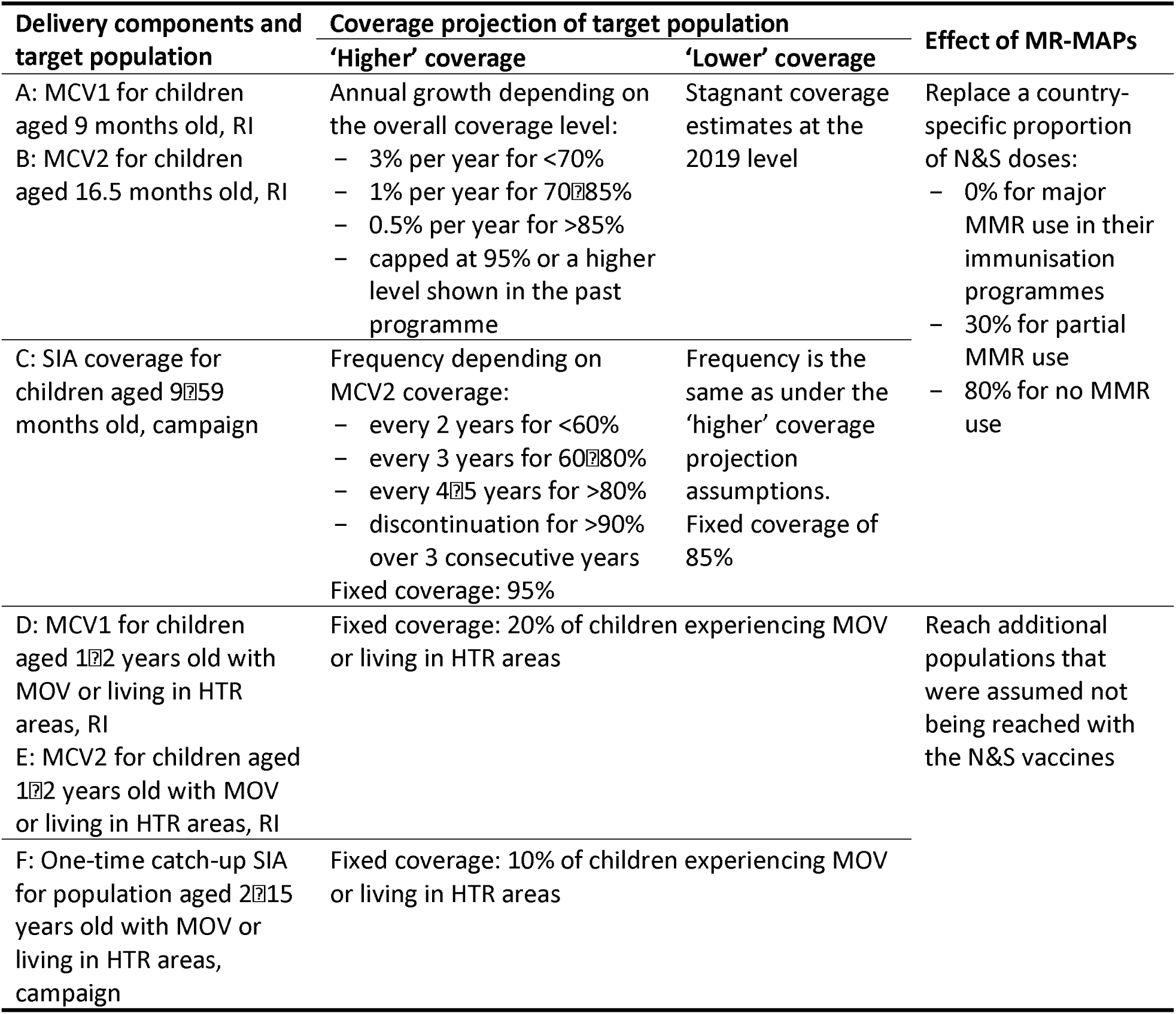
Assumptions for coverage forecasts and delivery components of measles vaccines. Measles vaccine delivery is modelled through six components (A to F) with different age and vaccination status of target populations, coverage projection assumptions, delivery approaches (RI or campaign), and dose presentations (N&S or MR-MAP). Details of the parameters and data sources used in shaping these assumptions are included in the demand forecast analysis by Ko et al.^27^. Introducing MR-MAPs was assumed to partially replace doses in the existing needle-based immunisation programmes with MR-MAPs (components A, B, and C) and provide additional MR-MAP doses to children with MOV or living in HTR areas (components D, E, and F). The level of replacement with MR-MAPs (market penetration) depends on the size of the different use cases for MR-MAPs and the characteristics of the measles and rubella programmes (inclusive of the use of MMR N&S vaccines) in each country. Abbreviations: HTR-hard-to-reach, MCV1-the first routine dose of measles-containing vaccine, MCV2-the second routine dose of measles-containing vaccine, MMR-measles-mumps-rubella, MOV-missed opportunities for vaccination, MR-MAP-measles-rubella microarray patch, N&S-needle and syringe, RI-routine immunisation, SIA-supplementary immunisation activity.

| Delivery components and target population | Coverage projection of target population |  | Effect of MR-MAPs |
| --- | --- | --- | --- |
|  | 'Higher' coverage | 'Lower' coverage |  |
| A: MCV1 for children aged 9 months old, RI<br>B: MCV2 for children aged 16.5 months old, RI | Annual growth depending on the overall coverage level:<br>– 3% per year for <70%<br>– 1% per year for 70–85%<br>– 0.5% per year for >85%<br>– capped at 95% or a higher level shown in the past programme | Stagnant coverage estimates at the 2019 level | Replace a country-specific proportion of N&S doses:<br>– 0% for major MMR use in their immunisation programmes<br>– 30% for partial MMR use<br>– 80% for no MMR use |
| C: SIA coverage for children aged 9–59 months old, campaign | Frequency depending on MCV2 coverage:<br>– every 2 years for <60%<br>– every 3 years for 60–80%<br>– every 4–5 years for >80%<br>– discontinuation for >90% over 3 consecutive years<br>Fixed coverage: 95% | Frequency is the same as under the 'higher' coverage projection assumptions. Fixed coverage of 85% |  |
| D: MCV1 for children aged 1–2 years old with MOV or living in HTR areas, RI<br>E: MCV2 for children aged 1–2 years old with MOV or living in HTR areas, RI | Fixed coverage: 20% of children experiencing MOV or living in HTR areas |  | Reach additional populations that were assumed not being reached with the N&S vaccines |
| F: One-time catch-up SIA for population aged 2–15 years old with MOV or living in HTR areas, campaign | Fixed coverage: 10% of children experiencing MOV or living in HTR areas |  |  |

**Table 2.** Unit costs (2020 USD) for measles vaccination and treatment by income level of countries. Unit costs per vaccine dose or per measles case are listed by income level and other factors specific to the type of costs. N&S vaccine procurement prices were extracted from the Market Information for Access Vaccine Purchase Database33 by taking the median prices over 201612020, while MR-MAP prices were estimated using the N&S prices plus potential increases in manufacturing costs (text A in online supplemental file S1). Wastage rates were specified by dose package and delivery approaches (RI or SIA) and applied to adjust vaccine price, as: procurement price / (1-wastage rate). Vaccine delivery costs are dependent on delivery approaches;^30^ for RI, marginal delivery cost increases with baseline coverage since more resources are required to reach the underserved populations. Costs for treating measles-related illness were extracted from countries with available data through a literature review (text B in online supplemental file S1). Abbreviations: MAPs-microarray patches, MI4A-Market Information for Access, MMR-measles-mumps-rubella, MR-measles-rubella, N&S-needle and syringe, RI-routine immunisation, USD-United States dollar.

| Cost type (per dose or per case) | Low income | Lower middle income | Upper middle income | Source and notes |
| --- | --- | --- | --- | --- |
| **N&S wastage-adjusted price, RI** | <u>Gavi-eligible countries, MR:</u><br>$0.73/(1-0.15) = \mathbf{0.86}$ (5-dose vial)<br>$0.66/(1-0.4) = \mathbf{1.10}$ (10-dose vial) | <u>Gavi, MR:</u><br>$0.82/(1-0.15) = \mathbf{0.96}$ (5-dose vial)<br>$0.66/(1-0.4) = \mathbf{1.10}$ (10-dose vial)<br><u>Non-Gavi, MR:</u><br>$0.87/(1-0.15) = \mathbf{1.02}$ (5-dose vial)<br>$0.70/(1-0.4) = \mathbf{1.17}$ (10-dose vial)<br><u>Non-Gavi, MMR:</u><br>$4.47/(1-0.01) = \mathbf{4.52}$ (2-dose vial) | <u>MR:</u><br>$2.37/(1-0.01) = \mathbf{2.39}$ (1-dose vial)<br>$0.69/(1-0.15) = \mathbf{0.81}$ (5-dose vial)*<br>$0.69/(1-0.4) = \mathbf{1.15}$ (10-dose vial)<br><u>MMR:</u><br>$4.00/(1-0.01) = \mathbf{4.04}$ (1-dose vial)*<br>$4.00/(1-0.01) = \mathbf{4.04}$ (2-dose vial)<br>$1.50/(1-0.15) = \mathbf{1.76}$ (5-dose vial)*<br>$1.50/(1-0.4) = \mathbf{2.50}$ (10-dose vial) | MI4A<sup>33</sup><br>1-dose vial: 1% wastage<br>2-dose vial: 1% wastage<br>5-dose vial: 15% wastage<br>10-dose vial: 40% wastage<br>*Prices from vials with a higher dose were used to ensure the inverse relationship between vial doses and costs. |
| **N&S wastage-adjusted price, campaign** | <u>Gavi, MR:</u><br>$0.73/(1-0.1) = \mathbf{0.81}$ (5-dose vial)<br>$0.66/(1-0.1) = \mathbf{0.73}$ (10-dose vial) | <u>Gavi, MR:</u><br>$0.82/(1-0.1) = \mathbf{0.91}$ (5-dose vial)<br>$0.66/(1-0.1) = \mathbf{0.73}$ (10-dose vial)<br><u>Non-Gavi, MR:</u><br>$0.87/(1-0.1) = \mathbf{0.97}$ (5-dose vial)<br>$0.70/(1-0.1) = \mathbf{0.78}$ (10-dose vial)<br><u>Non-Gavi, MMR:</u><br>$4.47/(1-0.01) = \mathbf{4.52}$ (2-dose vial) | <u>MR:</u><br>$2.37/(1-0.01) = \mathbf{2.39}$ (1-dose vial)<br>$0.69/(1-0.1) = \mathbf{0.77}$ (5-dose vial)*<br>$0.69/(1-0.1) = \mathbf{0.77}$ (10-dose vial)<br><u>MMR:</u><br>$4.00/(1-0.01) = \mathbf{4.04}$ (1-dose vial)*<br>$4.00/(1-0.01) = \mathbf{4.04}$ (2-dose vial)<br>$1.50/(1-0.1) = \mathbf{1.67}$ (5-dose vial)*<br>$1.50/(1-0.1) = \mathbf{1.67}$ (10-dose vial) | MI4A<sup>33</sup><br>1-dose vial: 1% wastage<br>2-dose vial: 1% wastage<br>5-dose vial: 10% wastage<br>10-dose vial: 10% wastage<br>*Prices from vials with a higher dose were used to ensure the inverse relationship between vial doses and costs. |
| **MR-MAP**<br>**wastage-adjusted**<br>**price, RI and**<br>**campaign** | 1.29/(1-0.01) = **1.30** (lower)<br>2.92/(1-0.01) = **2.95** (upper) | <u>Gavi:</u><br>1.29/(1-0.01) = **1.30** (lower)<br>2.92/(1-0.01) = **2.95** (upper)<br><u>Non-Gavi:</u><br>1.48/(1-0.01) = **1.49** (lower)<br>3.36/(1-0.01) = **3.39** (upper) | 2.63/(1-0.01) = **2.66** (lower)<br>5.20/(1-0.01) = **5.25** (upper) | 1-dose vial: 1% wastage<br>See text A in online<br>supplemental file S1 |
| **Vaccine delivery**<br>**cost, RI** | 2.02 plus<br>0.071 and 0.148 per 1% coverage<br>increase for baseline coverage<br><80% and >=80% | 5.11 plus<br>0.177 and 0.272 per 1% coverage<br>increase for baseline coverage<br><80% and >=80% | 6.45 | Levin et al.<sup>30</sup><br>Baseline coverage is set as<br>2020 projected coverage. |
| **Vaccine delivery**<br>**cost, campaign** | 0.91 | 1.89 | 1.97 | Levin et al.<sup>30</sup> |
| **Measles**<br>**treatment cost** | 10.9 | 120 | 235 | See text B in online<br>supplemental file S1 |

### MR-MAPs introduction

We modelled the effect of MR-MAPs introduction in partially replacing N&S doses in existing immunisation programmes (components A, B, and C in table 1) and reaching additional underserved populations for measles vaccination (components D, E, and F). Following the assumptions in the demand forecast analysis,^27^ the replacement of N&S doses with MR-MAP doses depends on the different MR-MAPs use cases and on country-specific groupings reflective of the characteristics of their measles and rubella immunisation programmes (table A in online supplemental file S1). In countries where measles-mumps-rubella (MMR) vaccines are widely adopted in their existing immunisation programmes, no MR-MAP doses are anticipated to be used, as implementing separate monovalent mumps vaccines with MR-MAPs is less programmatically feasible. In countries which only partially offer or do not use MMR vaccines, 30% or 80% of the total MCV doses will be replaced with MR-MAPs. In addition, with improved product characteristics, MR-MAPs are assumed to reach extra populations living in hard-to-reach areas and children with missed opportunities for vaccination, with a coverage of 20% through RI activities (delivery components D & E) and 10% through one-off campaigns (component F) in the targeted populations. The hard-to-reach populations include those in urban slums, security compromised, humanitarian settings, and remote/rural areas, while 2% of children under 2 years old were assumed to experience missed opportunities for measles vaccination.^27^

Two introduction strategies for MR-MAPs across countries were evaluated according to multiple factors, such as vaccine introduction history, disease burden, and funding for immunisation programmes.^27^ Under ‘sequential’ introduction, countries introduce MR-MAPs sequentially over the years between 2030⍰2040; those with higher measles and rubella burden and better operational and financial states for new vaccine introductions will adopt MR-MAPs in earlier years. Alternatively, ‘accelerated’ introduction allows countries with the greatest need, based on their MCV1 coverage and disease burden only, to be prioritised for MR-MAPs introduction. In figure 1, we present coverage forecasts under the sequential and accelerated strategies for MR-MAPs introduction, higher and lower coverage projection assumptions, and different delivery components (table 1) in the Democratic Republic of the Congo. The coverage forecasts for all 70 LMICs are included in online supplemental file S2.

**Figure 1.**
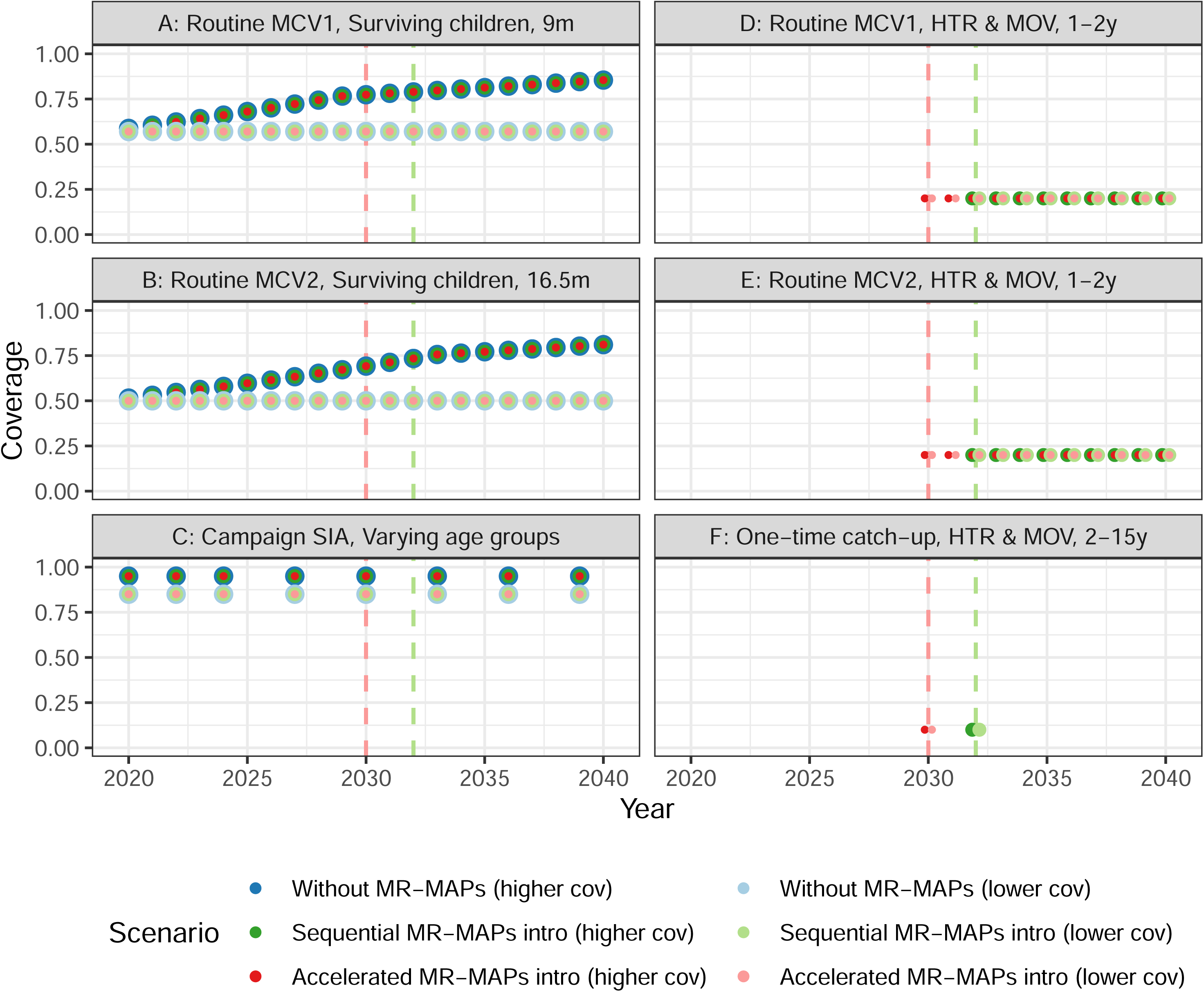
Coverage forecasts of measles vaccination by delivery components in the Democratic Republic of the Congo. Annual measles vaccine coverage forecasts through delivery components A⍰F (Table 1) are presented for the Democratic Republic of the Congo over 2020⍰2040. Circles with different colours show the coverage by scenarios, and vertical dashed lines represent the introduction years under the sequential (green) and accelerated (red) introduction. For components A⍰C, coverage forecasts for the existing immunisation programmes (blue) are the same regardless of MR-MAPs introduction but different for higher (darker colours) and lower (lighter colours) coverage projection assumptions. These coverage forecasts refer to the proportions of the total population in the corresponding age groups. For components D⍰F, the coverage forecasts refer to the proportions of children experiencing MOV or living in HTR areas that receive additional MR-MAP doses, and fixed coverage is assumed. Abbreviations: HTR-hard-to-reach, MCV1-the first routine dose of measles-containing vaccine, MCV2-the second routine dose of measles-containing vaccine, MOV-missed opportunities for vaccination, MR-MAP-measles-rubella microarray patch.

### Economic costs

We took a health provider perspective and ingredients approach in estimating the costs under different MR-MAPs scenarios. Table 1 shows unit costs of measles vaccine and treatment by country income level, with all values inflated to 2020 United States Dollar (USD) using country-specific Gross Domestic Product deflators.^28 29^ For each scenario, the total cost (treatment cost and vaccination cost) at year *t* in country *i* is denoted as:

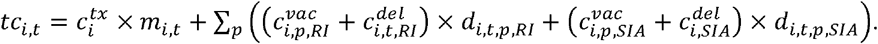

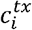represents the treatment cost for each measles case, extrapolated from data for countries at the same income level in the literature.^30-32^ *p* indicates different vaccine types characterised by dose presentation, vial size, and valent type. 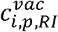 and 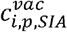 represent the vaccine procurement prices per dose delivered through RI and SIA, with an adjustment of wastage rates specific to the vaccine types. The procurement prices for N&S vaccines were obtained from the Market Information for Access Vaccine Purchase Database over 2016⍰2020.^33^ Taking the average price of MR N&S vaccines as the baseline, we estimated a potential range of prices for MR-MAPs by country income group and eligibility to receive Gavi funds. The lower MR-MAP price reflected the increased manufacturing cost per dose for a single-dose vial compared to a multiple-dose vial, using the price data of hepatitis B vaccines as a proxy. The difference in the cost for a pre-filled syringe compared to a single-dose vial is considered in estimating the upper MR-MAP price, based on the price information from pneumococcal conjugate vaccines (see Text A in online supplemental file S1 for details). 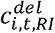 and 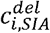 represent the vaccine delivery costs per dose for RI and SIA by income level. The marginal cost of delivering one routine MCV1 or MCV2 dose was assumed to increase with country-specific coverage forecasts, as higher coverage levels require extra resources for unvaccinated children having the least access to immunisation services.^30^ Finally, we multiplied these unit costs by the corresponding DynaMICE model estimates of *m*_*i,t*_ (the number of measles cases) and *d*_*i,t,p,RI*_ and *d*_*i,t,p,SIA*_ (the numbers of doses administered through RI and SIAs).

### Cost-effectiveness

Using DynaMICE, we estimated cases, deaths, disability-adjusted life years (DALYs), and economic costs under the sequential and accelerated MR-MAPs introduction and the baseline scenario without MR-MAPs. All the health and cost impact estimates were obtained at the country level and also aggregated by the World Bank income level.^18^ For each country and income group, we calculated the incremental cost-effectiveness ratio (ICER) by dividing the total incremental costs by the total averted DALYs over 2030⍰2040 between the MR-MAPs introduction scenarios and the baseline scenario. In the main analysis, a discounting rate of 3% was applied equally for costs and health outcomes. We also evaluated the ICERs under differential discounting, where only cost estimates were discounted at 3%, and health outcomes were not discounted, as recommended in the WHO guidelines for economic evaluation of immunisation programmes.^34^ Since GDP-based thresholds are no longer recommended by WHO,^35^ we instead used the cost-effectiveness thresholds estimated based on empirical data on the opportunity costs of healthcare expenditure^36 37^ to determine the cost-effectiveness of introducing MR-MAPs at the country level. As for the income-level ICERs, the thresholds were derived from population-weighted health opportunity costs in countries of the corresponding income groups (figure A in online supplemental file S1). In addition, we calculated the maximum MR-MAP procurement price that each country or income setting could afford to pay for the MR-MAPs introduction while ensuring the introduction is cost-effective (compared with country-specific thresholds based on health opportunity costs). This price calculation was conducted with the assumption that the health gains during 2030⍰2040 from preventing measles illness, vaccine wastage rates, and all the other types of costs are held fixed under equal discounting.

## Results

### Health impact of MR-MAPs introduction

Introduction of MR-MAPs would result in measles burden reductions across all country income levels and assumptions about coverage projections (figure 2). Under the higher coverage projection, 21.3 million cases, 197 thousand deaths, and 12.4 million DALYs due to measles are projected to occur over 2030⍰2040 across all the study countries if MR-MAPs are not available. Sequential introduction of MR-MAPs will result in 14.4 million measles cases, 145 thousand deaths, and 9.01 million DALYs, as accelerated introduction will result in 13.9 million measles cases, 139 thousand deaths, and 8.69 million DALYs. Countries in the lower-middle-income group contribute most to the global burden. The scale of the cumulative burden under the lower coverage projection is larger than the higher coverage projection, while the relative burden trends between different introduction strategies are similar. Under the lower coverage projection, there are estimated 106 million measles cases, 1.15 million deaths, and 74.0 million DALYs if MR-MAPs are not available, and 67.1 million measles cases, 750 thousand deaths, and 47.7 million DALYs with accelerated MR-MAPs introduction. Table 3 shows how MR-MAPs introduction will reduce the total measles burden in 70 LMICs over 2030⍰2040 by 27%⍰37%. At the global level, the relative health impact of introducing MR-MAPs is similar between the ‘higher’ and ‘lower’ coverage projection assumptions. However, the absolute impact varies, with 6.96⍰7.45 million and 31.3⍰38.6 million measles cases being averted by MR-MAP introduction under the higher and lower coverage projection assumptions, respectively. Overall, the accelerated introduction of MR-MAPs in key countries is projected to be more effective in reducing measles burden than the sequential introduction, since those countries with higher burden are more likely to benefit from the early introduction of MR-MAPs. For individual countries, introducing MR-MAPs will result in reduction of health burden over 2030⍰2040, although some variation remains within the same income-group level. We further analysed the exceptional countries and scenarios showing increased health burden following the MR-MAPs introduction and found health benefits of MR-MAPs introduction with an extended assessment period. This suggested that a longer time horizon would be needed to observe the complete health impacts of introducing MR-MAPs.

**Table 3.**
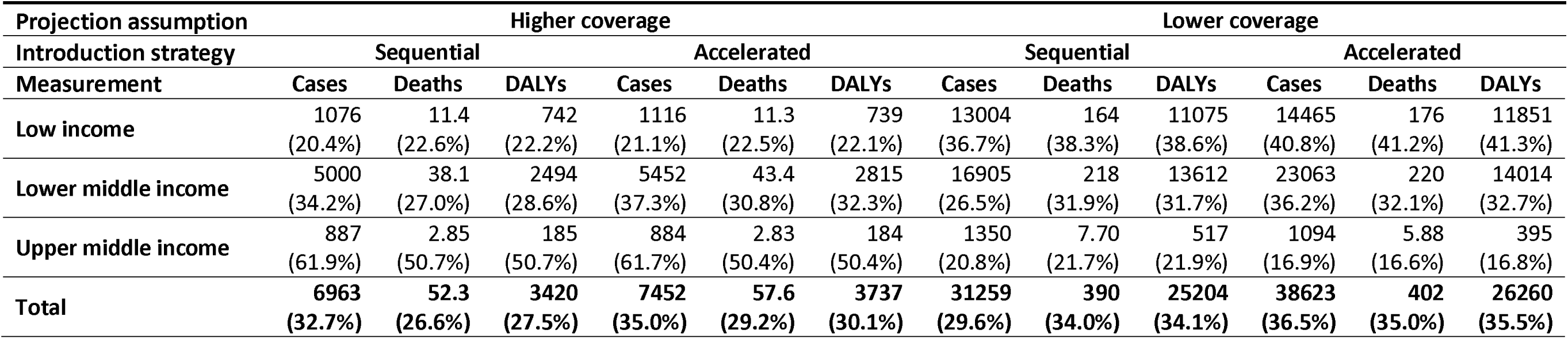
Averted measles burden (in thousands) following the introduction of MR-MAPs. Numbers represent the absolute cases, deaths, and DALYs averted in thousands. Percentages in the brackets show the relative burden reduction compared to the scenarios without MR-MAPs. Abbreviations: DALY-disability-adjusted life year, MR-MAP-measles-rubella microarray patch.

**Figure 2.**
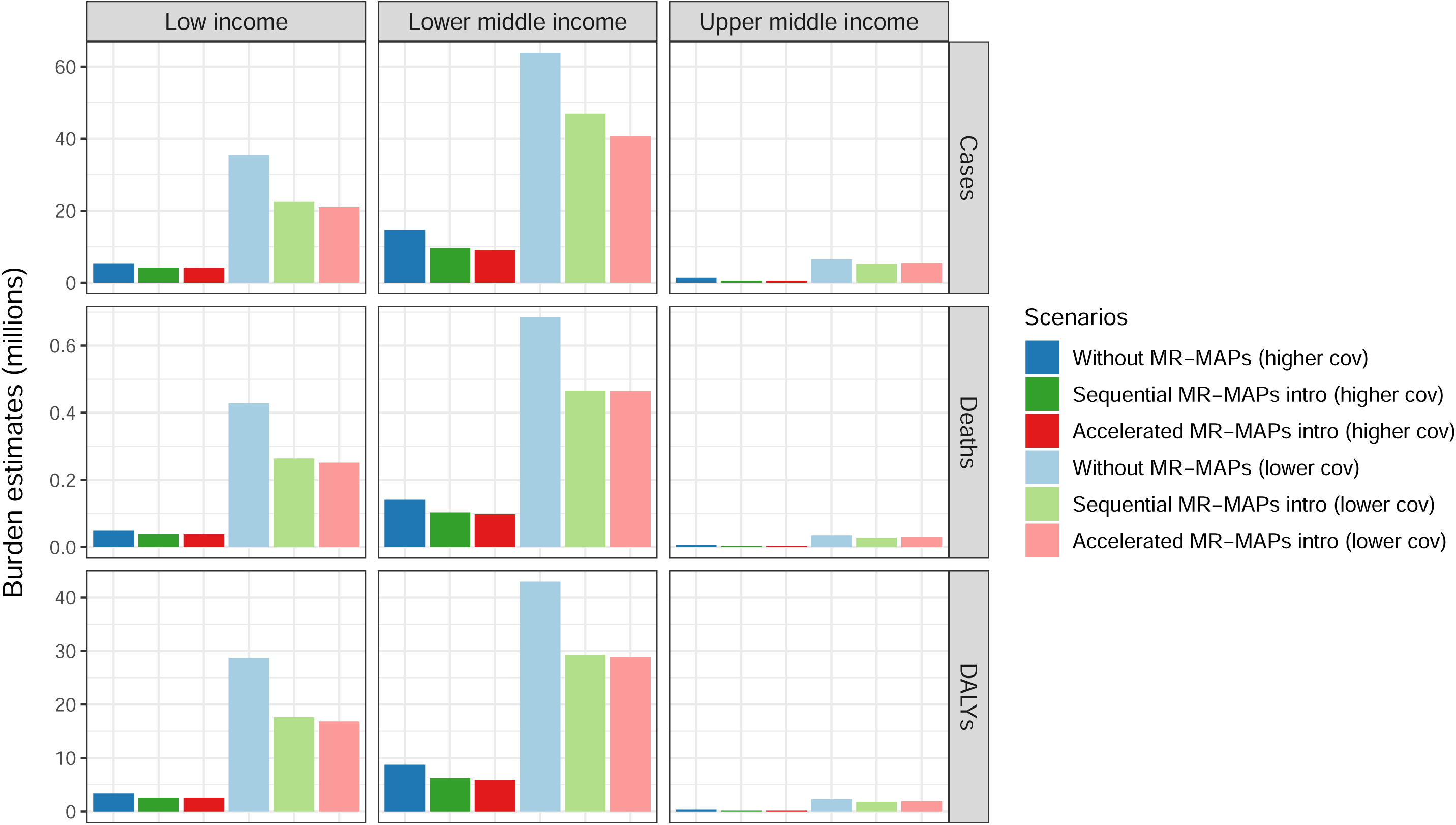
Cumulative measles burden (in millions) by coverage projections and income levels. Bars represent the total estimated measles cases, deaths, and DALYs in millions over 2030⍰2040 by income levels, with darker and lighter colours showing the results under higher and lower coverage assumptions, respectively. Abbreviations: DALY-disability-adjusted life year, MAP-microarray patch, MR-measles-rubella.

### Economic cost of MR-MAPs introduction

Figure 3 illustrates the breakdown of incremental costs following MR-MAPs introduction, compared to the baseline scenario without MR-MAPs. The introduction of MR-MAPs will reduce costs for the existing immunisation service based on N&S vaccines, which will be partly replaced with MR-MAP doses. However, the reduction will not exceed the increase in costs of purchasing and delivering MR-MAPs, since introducing MR-MAPs will deliver extra doses to children experiencing missed opportunities for vaccination and living in hard-to-reach areas (delivery components DCF in table 1). Savings from treating measles-associated illness after the MR-MAPs introduction were seen in all country income groups but relatively small compared to other cost types in the low-income setting. Overall, Introducing MR-MAPs will be cost-saving in the lower-middle-income and upper-middle-income settings provided at a lower MR-MAP procurement price under the lower coverage projection assumption (table C in online supplemental file S1) due to avoiding the high costs in these countries associated with measles treatment. In the low-income setting, the total incremental costs will increase across all the scenarios for evaluation. Unlike the MR-MAP price, the introduction strategies had little implications in the scale and direction of incremental costs. At the country level, the incremental costs following the introduction of MR-MAPs show a large variation within the same income group, as a result of country heterogeneities in dose demand, measles burden, and vaccine procurement costs.

**Figure 3.**
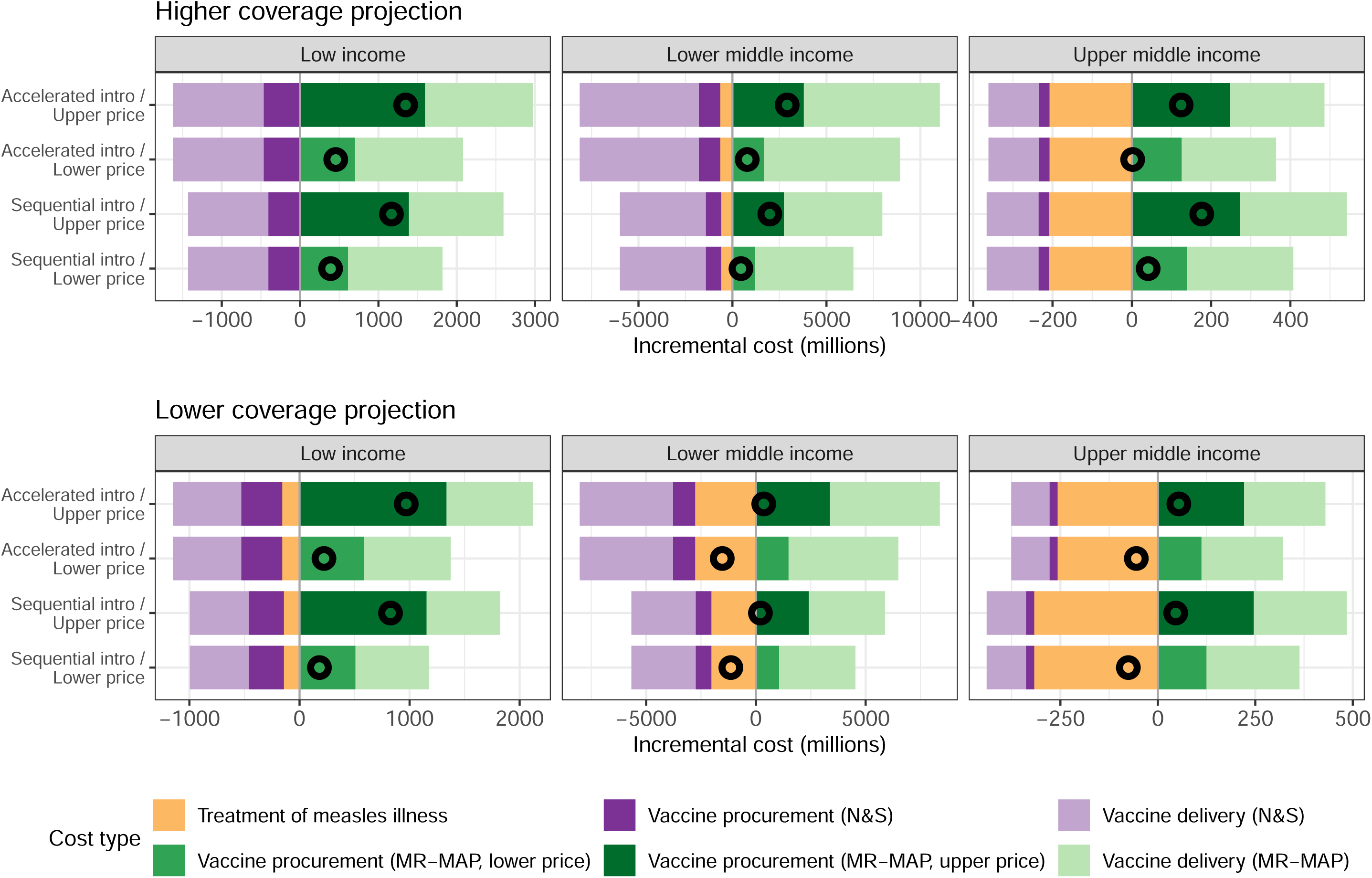
Breakdown of incremental costs following MR-MAPs introduction. The upper and lower panels show the incremental costs under higher and lower coverage projection assumptions, respectively. For each horizontal bar, incremental costs of measles treatment, vaccine procurement, and vaccine delivery are stacked and denoted in different colours. Hollow circles represent overall incremental costs. The vertical line is set at zero; to its left, negative values (purple and yellow bars) indicate savings following the MR-MAPs introduction, while to its right, positive values (green bars) indicate increased costs. Abbreviations: DALY-disability-adjusted life year, MR-MAP-measles-rubella microarray patch.

### Cost-effectiveness of MR-MAPs introduction

Figure 4 shows the income-group level ICERs of introducing MR-MAPs by different assumptions for discounting, coverage projection, introduction strategy, and MR-MAP price. The ICERs lie between 10.6⍰1850 USD in the low-income group, -108⍰1100 USD in the lower-middle-income group, and -134⍰1210 USD in the upper-middle-income group. Negative ICER values corresponding to settings where it is cost-saving to introduce MR-MAPs, are mostly seen in the upper-middle-income group with a lower MR-MAP price. In the lower-middle-income group, introducing MR-MAPs would be cost-effective under the lower coverage projection. Under the higher coverage projection, it would not be cost-effective with an upper MR-MAP price using equal discounting but cost-effective using differential discounting. In the low-income group, introducing MR-MAPs would only be cost-effective under the lower coverage projection, regardless of the set price for MR-MAPs. Assumptions about the introduction strategy resulted in smaller variations in the ICER estimates among low-income, lower-middle-income, and upper-middle-income country groups. Evaluating at the country level, we found similarity to the income-level analysis in factors that affect the cost-effectiveness of introducing MR-MAPs. MR-MAPs introduction would be less likely to be cost-effective in countries with a lower income, despite wide variation within each income group (figure B in online supplemental file S1). With a 3% discount rate for both health and cost impacts, the introduction of MR-MAPs over 2030⍰2040 is considered cost-effective in 26⍰81% and 16⍰61% of analysed countries under a lower and upper price of MR-MAPs, respectively (table 4). Alternatively, when applying differential discounting rates, MR-MAPs introduction would be cost-effective in 30⍰81% and 19⍰71% of countries under a lower and higher assumed price, respectively (table D in online supplemental file S1). The assumptions about coverage projection had the greatest influence on the cost-effectiveness of the MR-MAPs introduction. Meanwhile, the procurement cost for MR-MAPs was influential on its cost-effectiveness in lower-middle-income countries. Introducing MR-MAPs will be most cost-effective when provided through accelerated introduction with a lower MR-MAP price.

**Table 4.** Number of countries where introducing MR-MAPs is cost-effective. Introducing MR-MAPs is considered to be cost-effective if the country-specific ICER is below the country-specific threshold, under a 3% annual discount rate on both cost and health benefits. Abbreviations: ICER-incremental cost-effectiveness ratio, MR-MAP-measles-rubella microarray patch.

| Projection assumption | Higher coverage |  |  |  | Lower coverage |  |  |  |
| --- | --- | --- | --- | --- | --- | --- | --- | --- |
| Introduction strategy | Sequential |  | Accelerated |  | Sequential |  | Accelerated |  |
| MR-MAP price | Lower | Upper | Lower | Upper | Lower | Upper | Lower | Upper |
| Low income | 3/20 | 0/20 | 3/20 | 0/20 | 17/20 | 9/20 | 18/20 | 10/20 |
| Lower middle income | 10/35 | 6/35 | 9/35 | 5/35 | 25/35 | 20/35 | 28/35 | 22/35 |
| Upper middle income | 5/15 | 5/15 | 7/15 | 6/15 | 11/15 | 11/15 | 11/15 | 11/15 |
| Total | 18/70 | 11/70 | 19/70 | 11/70 | 53/70 | 40/70 | 57/70 | 43/70 |

**Figure 4.**
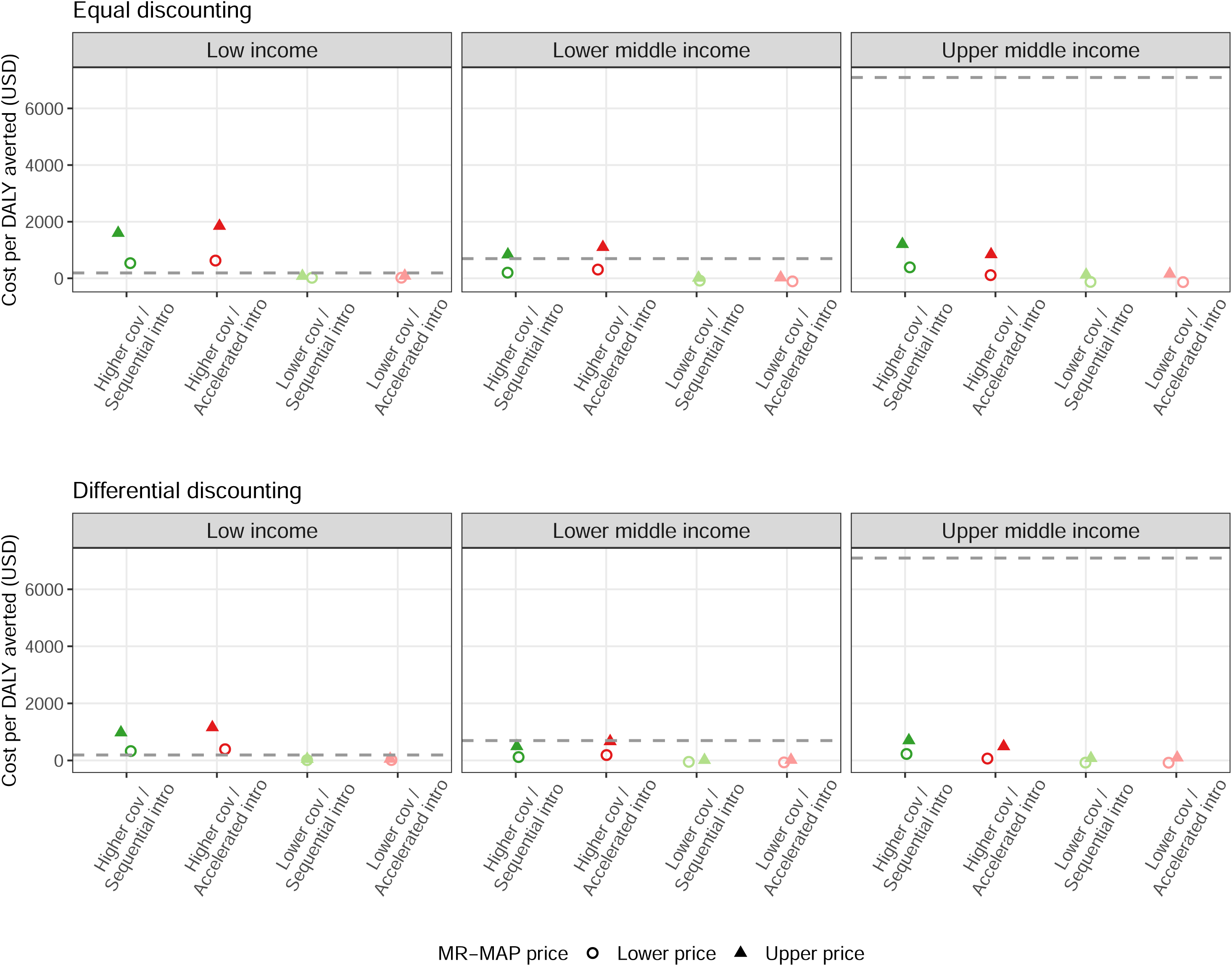
Incremental cost-effectiveness ratios of introducing MR-MAPs. Income-group level ICERs of introducing MR-MAPs are presented by discounting approaches. Equal discounting applies a 3% rate to both costs and health outcomes (averted DALYs), while differential discounting applies a 3% rate only to costs and no discounting of health outcomes. Circles and triangles denote the ICERs with a lower and upper MR-MAP price, respectively. Colours represent the sequential (green) and accelerated (red) introduction strategies and assumptions of higher (darker) and lower (lighter) coverage projections. Horizontal dashed lines indicate the cost-effectiveness thresholds at the income-group level; for scenarios with an ICER below the lines, it would be cost-effective to introduce MR-MAPs. Abbreviations: DALY-disability-adjusted life years, ICER-incremental cost-effectiveness ratio, MR-MAP-measles-rubella microarray patch.

The maximum per MR-MAP dose procurement prices (2020 USD) that ensure the cost-effectiveness of introducing MR-MAPs increase with the income-group level (table 5). The price thresholds for MR-MAPs are consistently higher compared to the corresponding prices for N&S vaccines (table 2). Under the lower coverage projection, the maximum MR-MAP prices are higher because of larger health benefits and savings from averting measles burden. In the low-income and lower-middle-income settings, the price thresholds for MR-MAPs were lower under the accelerated introduction strategy than the sequential strategy. While the accelerated introduction will bring greater health benefits, per-dose vaccine delivery cost will also increase with higher coverage, resulting in overall reduction in cost-effectiveness, particularly several years after the MR-MAPs introduction. As seen in the model-based health and cost impact estimates, the threshold prices across countries also result in great variability. In some countries, introducing MR-MAPs was found not to be cost-effective even if the procurement of MR-MAPs is zero price. These countries mostly have high measles vaccine coverage forecasts and only contributed to a small proportion (8%) of the total measles burden in the 70 LMICs between 2030⍰2040. Moreover, our price threshold analysis did not consider the potential reduction in delivery costs for MR-MAPs, which could improve the cost-effectiveness of the MR-MAPs introduction.

**Table 5.** Wastage-adjusted MR-MAP price thresholds (2020 USD) for introducing MR-MAPs to be cost-effective. Numbers represent the wastage-adjusted price thresholds for introducing MR-MAP to be cost-effective at the income-group and the country level by different coverage projection assumptions and introduction strategies. At the country level, the ranges of price thresholds are presented, with n in the brackets denoting the number of countries except for those where introducing MR-MAPs will not be cost-effective even if the procurement of MR-MAPs is at zero cost. The price thresholds were calculated while the health burden estimates, vaccine wastage rates, and other cost inputs were assumed fixed. If an MR-MAP dose is provided at a procurement price above the threshold, it implies that introducing MR-MAPs would not be cost-effective. Abbreviation: MR-MAP-measles-rubella microarray patch.

| Projection assumption | Higher coverage |  | Lower coverage |  |
| --- | --- | --- | --- | --- |
| Introduction strategy | Sequential | Accelerated | Sequential | Accelerated |
| **Low income** |  |  |  |  |
| Income-group level | 0.764 | 0.709 | 5.89 | 5.47 |
| Country level | 0.123–2.32 (n = 19) | 0.041–2.51 (n = 19) | 0.397–18.1 (n = 19) | 0.586–18.1 (n = 19) |
| **Lower middle income** |  |  |  |  |
| Income-group level | 2.67 | 2.17 | 14.7 | 11.6 |
| Country level | 0.012–37.9 (n = 25) | 0.051–40.7 (n = 27) | 0.086–39.6 (n = 31) | 0.572–72.0 (n = 29) |
| **Upper middle income** |  |  |  |  |
| Income-group level | 23.9 | 27.4 | 76.6 | 67.2 |
| Country level | 0.658–353 (n = 10) | 0.635–389 (n = 9) | 0.586–3310 (n = 12) | 0.579–1730 (n = 12) |

## Discussion

Our modelling analysis suggests that the introduction of MR-MAPs will bring substantial health benefits over 2030⍰2040 and is likely to be cost-effective in 16⍰81% of LMICs, depending on the assumptions used. The strategy of accelerating MR-MAPs introduction in high-burden countries generates the largest burden reduction compared to sequential introduction globally, despite requiring delayed introduction in lower-burden countries. Assumptions about the underlying MCV coverage growth in the future have a great influence on the magnitude of health impacts from introducing MR-MAPs, where the lower coverage projection resulted in larger numbers of averted cases, deaths, and DALYs. Additionally, among the different types of costs, the cost for treating measles illness has a key role in the total expenditure and savings on treatment costs would surpass the incremental costs for the MR-MAPs introduction, especially in countries with a higher income. For each country, the cost-effectiveness of introducing MR-MAPs will largely depend on the procurement price of a MR-MAP dose.

To our knowledge, our study is the first cost-effectiveness assessment at a global scale on the potential impact of the MR-MAP introduction. A previous study by Adhikari et al. found that replacing traditional N&Ss with MAPs for measles immunisation is cost-effective in a hypothetical population, due to cost reductions in cold chains, personnel, injection equipment, and needle disposal.^38^ However, the detailed cost changes following the introduction of MR-MAPs would need more empirical research and data to estimate.

Additionally, Adhikari et al. assumed no changes in the total administered doses,^38^ while we focused on the possible integration of MR-MAPs into existing immunisation programmes, with partial penetration of the N&S market and additional vaccine delivery to the populations living in hard-to-reach areas and experiencing missed opportunities of vaccination. We considered both the cost and health aspects of MR-MAPs and included the health benefits of reaching previously under-immunised populations.

Assumptions about future coverage projections exert the greatest influence on the health impact of MR-MAPs among the assumptions examined in this study. The wide range between the lower and higher coverage projection assumptions aims to reflect the multiple sources of uncertainties around the future progress of national immunisation programmes and measles elimination efforts. The pessimistic lower coverage projection scenario may not capture worst-case situations such as those caused by funding instability, emerging diseases, natural disasters, and political conflicts. Such situations could cause MCV coverage not just to stagnate, but to drop below what has been achieved historically, as seen in the global coverage estimate that reduced from 84% in 2020 to 81% in 2021 following the COVID-19 pandemic.^5^ The role of MR-MAPs could be even more critical in measles immunisation when the future performance of immunisation programmes cannot meet the historical levels of coverage. In addition, our analysis shows a potentially substantial impact on measles burden reduction with a 10⍰20% increase in coverage from providing MR-MAPs to populations living in hard-to-reach areas and experiencing missed opportunities for vaccination. With further reach to these under-immunised populations, MR-MAPs would realise even greater benefits in reducing measles burden.

To estimate the incremental cost of MR-MAPs introduction, we varied vaccine procurement costs, delivery costs, and measles treatment costs by country income level. However, we assumed that delivery costs were the same for N&Ss and MR-MAPs, which may underestimate the potential of MR-MAPs to save costs from health personnel capacity and cold chain equipment.^38^ On the other hand, integration of MR-MAPs into existing immunisation programmes will require structural changes in staffing and operation. There are uncertainties in estimating the delivery costs of MR-MAPs. Collecting the cost data, even at an early stage of vaccine development and licensure, will be useful in informing the investment case of future introduction. Indirect costs were not systematically included in this study. Patient costs for travel and waiting time during vaccination visits were included in a few original data sources, but productivity costs around the treatment of measles-associated illness were not considered.^30^ From a societal perspective, including productivity loss would make the introduction of MR-MAPs more cost-effective. Although MR-MAPs introduction was not considered cost-effective in some upper-middle-income settings from the perspective of measles burden reduction, their ability to reach under-immunised populations may still drive the introduction in order to reach regional measles elimination goals earlier. Furthermore, the impact of an earlier national/regional measles elimination, with the respective savings including reduced future demand for measles vaccines, was not considered in the cost-effectiveness analysis and may further increase the cost-effectiveness of MR-MAPs introduction.

There are other limitations in the cost-effectiveness analysis. First, we estimated the health burden following MR-MAPs introduction based on country-level data and models, but some of the largest disparities in vaccine coverage exist at the subnational level such as in remote rural areas or border regions.^39^ We did not investigate whether subnational targeting of MR-MAPs may achieve most of the coverage improvements at reduced cost. Second, we assessed the health and economic benefits of measles burden reduction following the introduction of MR-MAPs but excluded the concurrent reduction of rubella burden. Taking into account the impact on rubella burden will make the introduction of MR-MAPs more cost-effective. Nonetheless, the additional benefits may not be substantial compared to the reduction in measles burden since rubella is less transmissible and typically less severe than measles. Third, measles control measures apart from vaccination, such as contact tracing, self-quarantine, and post-exposure prophylaxis, were not included in the epidemiological modelling. This may affect both the cost and burden estimates, particularly in countries close to elimination.^40^ However, their effect may be more limited in the high-burden countries that we examined. Finally, cost-effectiveness thresholds based on health opportunity costs may be useful in reflecting the value for money from a fixed budget perspective, but the methodology for this estimation has not yet been maturely developed, and only few countries have adopted these thresholds explicitly in decision-making for health policies.^41^ Other context-specific considerations may also be decisive factors in the funding and implementation of MR-MAPs, and WHO recommends against basing health investment decisions purely on comparison to a cost-effectiveness threshold.^35 42 43^

This early-phase cost-effectiveness analysis was conducted from the perspective of the funder, e.g. the country or pooled procurement donors such as Gavi. The examination of the return to the manufacturer on the investment needed for research and development before MR-MAPs enter the market, and the net present value of MR-MAPs are concurrently developed in the initial Full Value of Vaccines Assessment of MR-MAPs and will be reported (*DOI to be added once it is officially released*.). Our analysis of the cost-effectiveness, including the calculated price thresholds for MR-MAPs, may inform vaccine investment entities such as the Vaccine Innovation Prioritisation Strategy about strategies to increase manufacturers’ incentives to develop and market MR-MAPs, in order to achieve the projected health impact. While MR-MAPs are still in the early stage of clinical development, this analysis can provide initial but comprehensive evidence that policy makers can apply to understand the potential cost-effectiveness of MR-MAPs at the global and/or national level and consider the implementation of MR-MAPs in their longer-term measles vaccination strategies.

In conclusion, this study has shown that there are substantial health benefits from introducing MR-MAPs in LMICs, particularly if countries bearing greater measles burden can be prioritised, as evaluated in the scenario with accelerated introduction. In addition to the reduction in measles burden, MR-MAPs introduction would accelerate progress towards measles elimination through more effectively and equitably reaching underserved populations of children with missed opportunities for vaccination or living in hard-to-reach areas. Introducing MR-MAPs is also cost-effective in most LMICs, while external funding from donors could support the MR-MAPs introduction in countries where it is not cost-effective under fixed domestic budgets. Although several assumed features of MR-MAPs are still highly uncertain, this early-stage evaluation can help inform key data gaps in assessing the cost-effectiveness of MR-MAPs, including future MCV coverage assumptions, MR-MAP ability to reach zero-dose and under-immunised children, procurement prices, and country-specific costs for measles treatment and vaccine delivery. Addressing these data gaps will facilitate evidence generation for decision-making on MR-MAPs introductions in LMICs.

## Data Availability

Data and codes produced in the present are available upon reasonable request to the authors and will be made public in a GitHub repository once the manuscript is accepted for journal publication.

## Data availability statement

Relevant datasets and computer codes of this analysis are available from the corresponding author on reasonable request and will be made public in the GitHub repository once the manuscript is accepted for journal publication.

## Ethics statements

### Patient consent for publication

Not applicable

### Ethics approval

Not applicable

## Acknowledgements

We thank Simon Procter at London School of Hygiene and Tropical Medicine, Jessica Ochalek at York University, and Adam Soble, Thomas Cherian, and Carsten Mantel at MMGH Consulting GmbH for helpful discussions. We acknowledge the feedback from members of the External Advisory Group of the initial Full Vaccine Value Assessment (iFVVA) for MR-MAPs and the WHO Immunization and Vaccine-related Implementation Research Advisory Committee.

## Supplementary Files

**Online supplemental file S1. Supplementary methods and results**. Text A. Estimating the theoretical price ranges for MR-MAPs. Text B. Cost of treating a measles case. Table A. List of 70 countries and related inputs included in the study. Table B. DynaMICE model parameters. Table C. Incremental cost (in million USD) of introducing MR-MAPs by income groups, coverage assumptions, and the MR-MAP prices. Table D. Number of countries where introducing MR-MAPs is cost-effective under differential discounting. Figure A. Health opportunity costs in 70 low-and middle-income countries. Figure B. Net health benefits of introducing MR-MAPs in 70 countries under the equal discounting assumption.

**Online supplemental file S2. Coverage forecasts and delivery components in 70 countries**.

## References

1. Keja K, Chan C, Hayden G, et al. Expanded programme on immunization. World Health Stat Q 1988;41(2):59–63. [published Online First: 1988/01/01]

2. Li X, Mukandavire C, Cucunuba ZM, et al. Estimating the health impact of vaccination against ten pathogens in 98 low-income and middle-income countries from 2000 to 2030: a modelling study. Lancet 2021;397(10272):398–408. doi: 10.1016/S0140-6736(20)32657-X [published Online First: 2021/02/01]

3. World Health Organization. Measles vaccines: WHO position paper - April 2017. Weekly Epidemiological Record 2017;92(17):205–27. [published Online First: 2017/05/02]

4. Causey K, Fullman N, Sorensen RJD, et al. Estimating global and regional disruptions to routine childhood vaccine coverage during the COVID-19 pandemic in 2020: a modelling study. Lancet 2021 doi: 10.1016/S0140-6736(21)01337-4 [published Online First: 2021/07/18]

5. World Health Organization. Immunization data portal, 2021.

6. Forster AH, Witham K, Depelsenaire ACI, et al. Safety, tolerability, and immunogenicity of influenza vaccination with a high-density microarray patch: Results from a randomized, controlled phase I clinical trial. PLoS Med 2020;17(3):e1003024. doi: 10.1371/journal.pmed.1003024 [published Online First: 2020/03/18]

7. The Vaccine Innovation Prioritisation Strategy. VIPS phase 2 techinical note: Microarray patches, 2020.

8. Gavi, the Vaccine Alliance. Vaccine microarray patches (MAPs): public summary of the VIPS Alliance Action Plan. 2021

9. Richardson LC, Moss WJ. Measles and rubella microarray array patches to increase vaccination coverage and achieve measles and rubella elimination in Africa. Pan Afr Med J 2020;35(Suppl 1):3. doi: 10.11604/pamj.supp.2020.35.1.19753 [published Online First: 2020/05/07]

10. Hasso-Agopsowicz M, Crowcroft N, Biellik R, et al. Accelerating the Development of Measles and Rubella Microarray Patches to Eliminate Measles and Rubella: Recent Progress, Remaining Challenges. Front Public Health 2022;10:809675. doi: 10.3389/fpubh.2022.809675 [published Online First: 2022/03/22]

11. Peyraud N, Zehrung D, Jarrahian C, et al. Potential use of microarray patches for vaccine delivery in low- and middle-income countries. Vaccine 2019;37(32):4427–34. doi: 10.1016/j.vaccine.2019.03.035 [published Online First: 2019/07/03]

12. Joyce JC, Carroll TD, Collins ML, et al. A Microneedle Patch for Measles and Rubella Vaccination Is Immunogenic and Protective in Infant Rhesus Macaques. J Infect Dis 2018;218(1):124–32. doi: 10.1093/infdis/jiy139 [published Online First: 2018/04/28]

13. Measles and Rubella Vaccine Microneedle Patch Phase 1-2 Age De-escalation Trial 2020 [Available from: https://ClinicalTrials.gov/show/NCT04394689 accessed 1 May 2022.

14. Vaxxas Pty Ltd. First-in-human study to investigate the skin tolerability of micro-projection array patches coated with live attenuated measles and rubella vaccine in healthy adult volunteers: https://anzctr.org.au/Trial/Registration/TrialReview.aspx?ACTRN=12621000820808, 2021.

15. Newall AT, Beutels P, Tuffaha HW, et al. How can early stage economic evaluation help guide research for future vaccines? Vaccine 2022;40(2):175–77. doi: 10.1016/j.vaccine.2021.11.017 [published Online First: 2021/12/07]

16. World Health Organization, United Nations Children’s Fund. Measles-rubella microarray patch (MR–MAP) target product profile. Switzerland, 2019.

17. Hutubessy RCW, Lauer JA, Giersing B, et al. The Full Value of Vaccine Assessments (FVVA): A Framework to Assess and Communicate the Value of Vaccines for Investment and Introduction Decision Making. SSRN 2021 doi: https://dx.doi.org/10.2139/ssrn.3841999 [published Online First: May 7]

18. The World Bank. World Bank Country and Lending Groups.

19. Fu H, Abbas K, Klepac P, et al. Effect of evidence updates on key determinants of measles vaccination impact: a DynaMICE modelling study in ten high-burden countries. BMC Med 2021;19(1):281. doi: 10.1186/s12916-021-02157-4 [published Online First: 2021/11/18]

20. Prem K, van Zandvoort K, Klepac P, et al. Projecting contact matrices in 177 geographical regions: an update and comparison with empirical data for the COVID-19 era. Plos Comput Bio 2021;17(7):e1009098. doi: 10.1371/journal.pcbi.1009098 [published Online First: 2021/07/26]

21. Guerra FM, Bolotin S, Lim G, et al. The basic reproduction number (R0) of measles: a systematic review. Lancet Infect Dis 2017;17(12):e420–e28. doi: 10.1016/S1473-3099(17)30307-9 [published Online First: 2017/08/02]

22. Portnoy A, Jit M, Ferrari M, et al. Estimates of case-fatality ratios of measles in low-income and middle-income countries: a systematic review and modelling analysis. Lancet Glob Health 2019;7(4):e472–e81. doi: 10.1016/S2214-109X(18)30537-0 [published Online First: 2019/02/25]

23. Cata-Preta BO, Santos TM, Mengistu T, et al. Zero-dose children and the immunisation cascade: Understanding immunisation pathways in low and middle-income countries. Vaccine 2021;39(32):4564–70. doi: 10.1016/j.vaccine.2021.02.072 [published Online First: 2021/03/22]

24. Hughes SL, Bolotin S, Khan S, et al. The effect of time since measles vaccination and age at first dose on measles vaccine effectiveness - A systematic review. Vaccine 2020;38(3):460–69. doi: 10.1016/j.vaccine.2019.10.090 [published Online First: 2019/11/17]

25. Sudfeld CR, Navar AM, Halsey NA. Effectiveness of measles vaccination and vitamin A treatment. Int J Epidemiol 2010;39 Suppl 1:i48-55. doi: 10.1093/ije/dyq021 [published Online First: 2010/04/02]

26. World Health Organization. Global market study: measles-containing vacclines (MCV), 2020.

27. Ko M, Malvoti S, Cherian T, et al. Estimating the future global dose demand for Measles-Rubella microarray patches. medRxiv 2022:2022.08.11.22278665. doi: 10.1101/2022.08.11.22278665

28. The World Bank. GDP deflator (base year varies by country). 1960-2020.

29. Turner HC, Lauer JA, Tran BX, et al. Adjusting for Inflation and Currency Changes Within Health Economic Studies. Value Health 2019;22(9):1026–32. doi: 10.1016/j.jval.2019.03.021 [published Online First: 2019/06/14]

30. Levin A, Burgess C, Garrison LP, Jr., et al. Global eradication of measles: an epidemiologic and economic evaluation. J Infect Dis 2011;204 Suppl 1:S98-106. doi: 10.1093/infdis/jir096 [published Online First: 2011/06/17]

31. Hyle EP, Fields NF, Fiebelkorn AP, et al. The Clinical Impact and Cost-effectiveness of Measles-Mumps-Rubella Vaccination to Prevent Measles Importations Among International Travelers From the United States. Clin Infect Dis 2019;69(2):306–15. doi: 10.1093/cid/ciy861 [published Online First: 2018/10/13]

32. Njau J, Janta D, Stanescu A, et al. Assessment of Economic Burden of Concurrent Measles and Rubella Outbreaks, Romania, 2011-2012. Emerging infectious diseases 2019;25(6):1101–09. doi: 10.3201/eid2506.180339 [published Online First: 2019/05/21]

33. World Health Organization. MI4A Vaccine Purchase Database.

34. World Health Organization. WHO guide for standardization of economic evaluations of immunization programmes. 2nd ed. Geneva, 2019.

35. Bertram MY, Lauer JA, De Joncheere K, et al. Cost-effectiveness thresholds: pros and cons. Bulletin of the World Health Organization 2016;94(12):925–30. doi: 10.2471/BLT.15.164418 [published Online First: 2016/12/21]

36. Lomas J, Claxton K, Ochalek J. Accounting for country- and time-specific values in the economic evaluation of health-related projects relevant to low- and middle-income countries. Health Policy Plan 2022;37(1):45–54. doi: 10.1093/heapol/czab104 [published Online First: 2021/08/20]

37. Ochalek J, Lomas J. Reflecting the Health Opportunity Costs of Funding Decisions Within Value Frameworks: Initial Estimates and the Need for Further Research. Clin Ther 2020;42(1):44–59 e2. doi: 10.1016/j.clinthera.2019.12.002 [published Online First: 2020/01/21]

38. Adhikari BB, Goodson JL, Chu SY, et al. Assessing the Potential Cost-Effectiveness of Microneedle Patches in Childhood Measles Vaccination Programs: The Case for Further Research and Development. Drugs R D 2016;16(4):327–38. doi: 10.1007/s40268-016-0144-x [published Online First: 2016/10/04]

39. Local Burden of Disease Vaccine Coverage C. Mapping routine measles vaccination in low- and middle-income countries. Nature 2020 doi: 10.1038/s41586-020-03043-4 [published Online First: 2020/12/18]

40. Enanoria WT, Liu F, Zipprich J, et al. The Effect of Contact Investigations and Public Health Interventions in the Control and Prevention of Measles Transmission: A Simulation Study. PloS one 2016;11(12):e0167160. doi: 10.1371/journal.pone.0167160 [published Online First: 2016/12/13]

41. Thokala P, Ochalek J, Leech AA, et al. Cost-Effectiveness Thresholds: the Past, the Present and the Future. Pharmacoeconomics 2018;36(5):509–22. doi: 10.1007/s40273-017-0606-1 [published Online First: 2018/02/11]

42. World Health Organization. WHO Evidence Considerations for Vaccine Policy Development (ECVP): generic framework for vaccines/monoclonal antibodies in development. In: Immunization VaB, ed., 2022.

43. Kochhar S, Barreira D, Beattie P, et al. Building the concept for WHO Evidence Considerations for Vaccine Policy (ECVP): Tuberculosis vaccines intended for adults and adolescents as a test case. Vaccine 2022;40(12):1681–90. doi: 10.1016/j.vaccine.2021.10.062 [published Online First: 2022/02/16]

